## Supplementary material for "Early menarche and childbirth accelerate aging-related outcomes and age-related diseases: Evidence for antagonistic pleiotropy in humans": figs S1 to S3

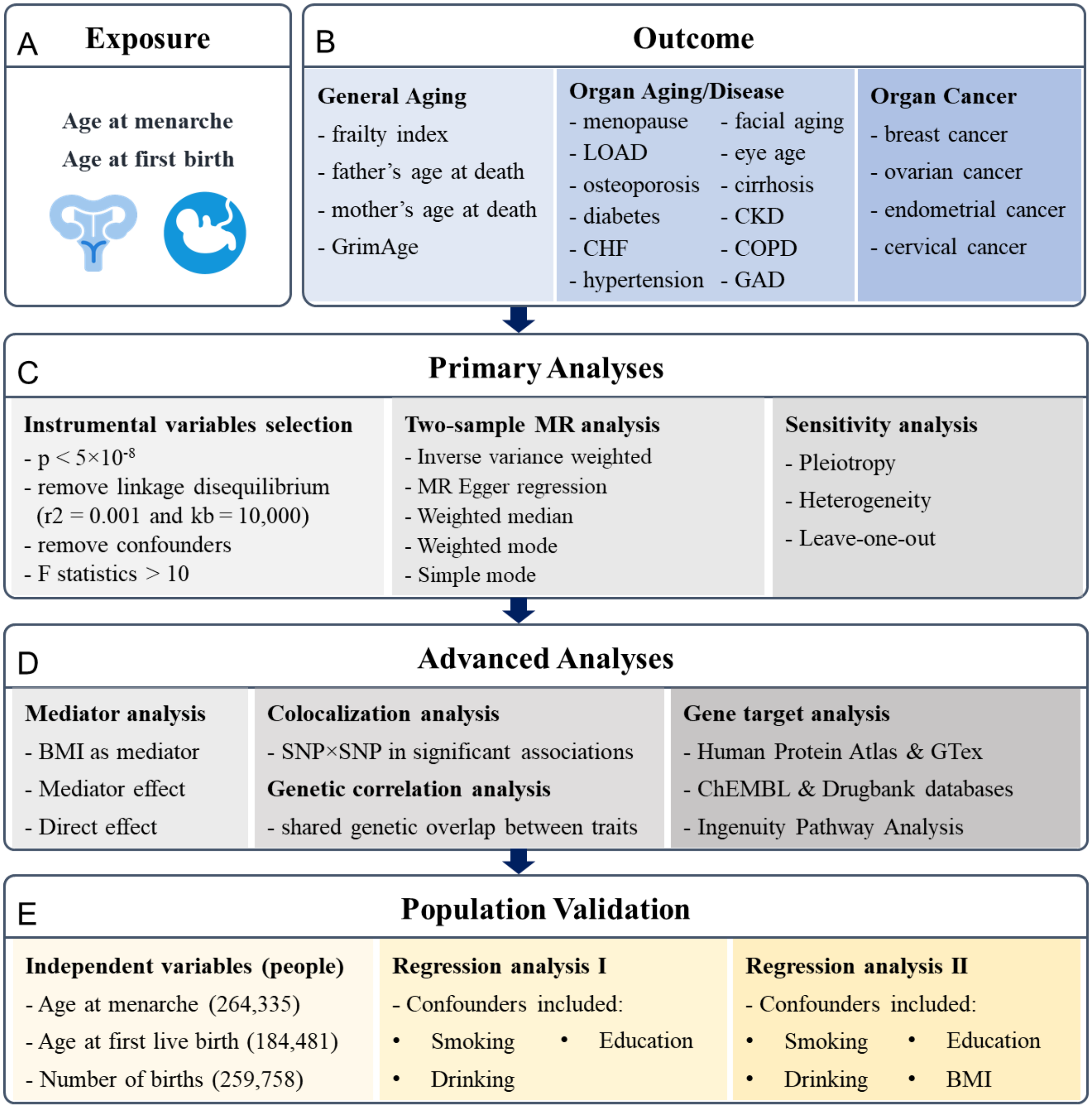


**Fig. S1. Research pipeline.** (**A**) and (**B**), all the traits for exposures and outcomes included in the analysis. Exposures include age at menarche and age at first birth. Outcomes include general aging, organ aging and diseases, and organ cancers. (**C**), the primary analyses for MR research, including instrumental variables selection, two-sample MR analysis, and sensitivity tests. (**D**), the advanced analyses include mediator analysis of BMI with two-step MR, colocalization analysis on SNP-SNP level, genetic correlation analysis based on LDSC, and gene target analysis based on RNA and protein expression analysis and Ingenuity Pathway Analysis. (**E**), the results are validated based on the UK Biobank with regression analysis. LOAD, late-onset Alzheimer's disease; CHF, chronic heart failure; CKD, chronic kidney disease; COPD, chronic obstructive pulmonary disease; GAD, gastrointestinal or abdominal disease.

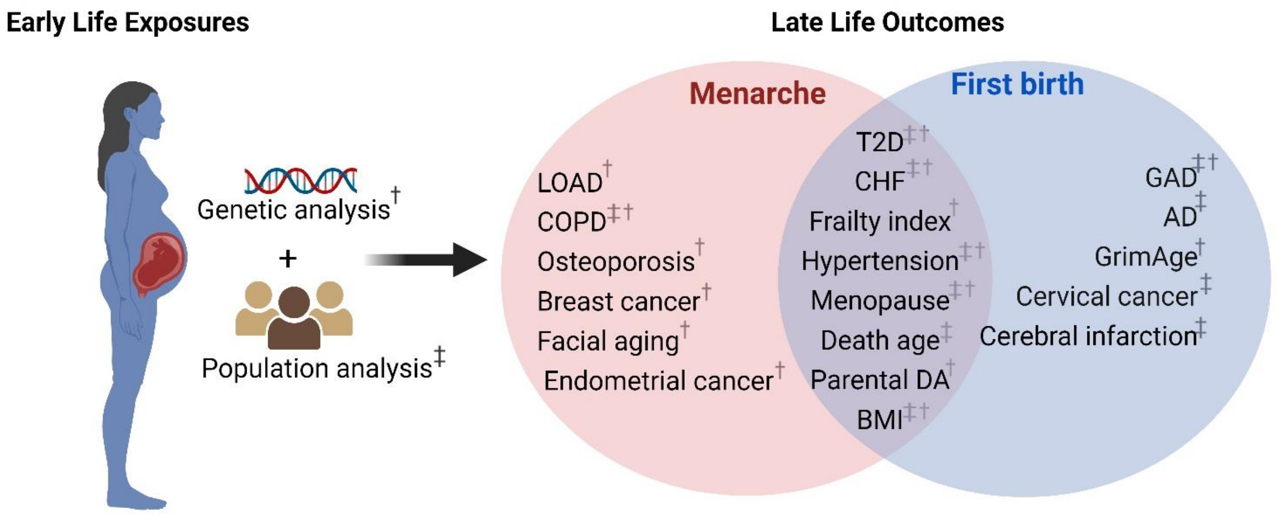


**Fig. S2. Schematic representation of early reproduction exposures and age-related outcomes.** †Significant age-related outcomes in MR analysis. ‡Significant age-related outcomes in UKBB cohorts. LOAD, late-onset Alzheimer's disease; COPD, chronic obstructive pulmonary diseases; T2D, type 2 diabetes; CHF, chronic heart failure; BMI, body mass index; Parental DA, parental death age; GAD, gastrointestinal and abdominal diseases; AD, Alzheimer's disease. The figure is created with BioRender.com.

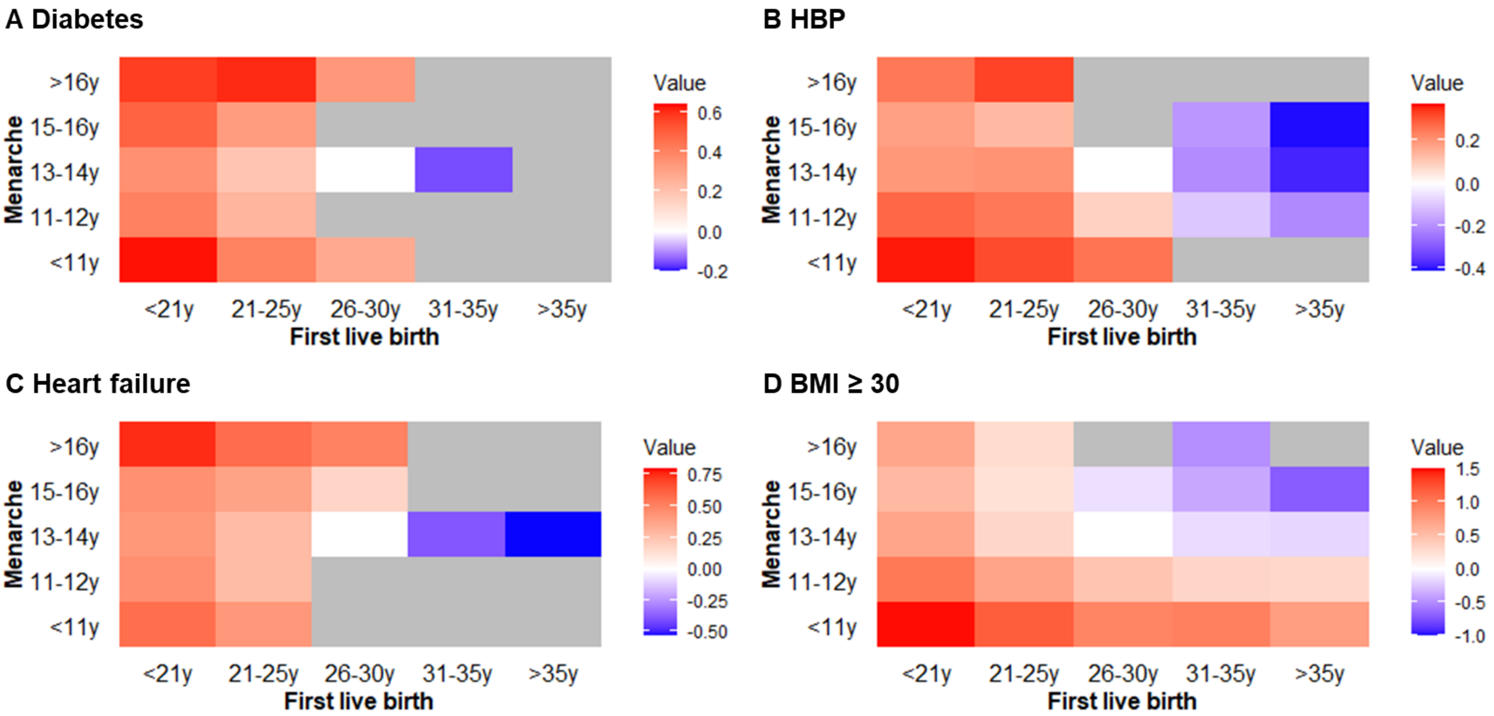


**Fig. S3. The combined effect of age at menarche and first live birth for different outcomes.** Females with menarche of "13-14y" and first birth of "26-30y" were set as the base line group in the analysis. Red block means higher risk. Blue block means lower risk. White block means baseline. Grey block means no significant difference. HBP, high blood pressure; BMI, body mass index.
